# Large Language Model - Enhanced Decision Tree Framework for Identifying Multiple Sclerosis Diagnoses from Clinical Documentation

**DOI:** 10.64898/2026.07.14.26357416

**Authors:** Shruthi Venkatesh, Marisa DelSignore, Xizhi Wu, Michele Morris, Wesley T. Kerr, Shyam Visweswaran, Yanshan Wang, Zongqi Xia

## Abstract

**Background.:** Early diagnosis and intervention are crucial in multiple sclerosis (**MS**), yet diagnostic delays are common. Large language models (**LLMs**) such as generative pre-trained transformers (**GPTs**) may help streamline diagnostic workflows by extracting MS diagnostic signals from clinical notes.

**Objective.:** To derive MS diagnosis status from the first neurology note using a computable algorithm based on the 2017 McDonald criteria and applying GPT-4 for node-level reasoning within a structured decision framework.

**Methods.:** We analyzed first neurology notes from 125 randomly selected patients (including those with MS, related disorders, and controls) enrolled in a clinic cohort between 2017 and 2023. We included the clinical history and diagnostic testing sections but redacted the assessment and plan. We converted the 2017 McDonald criteria into a decision tree and provided expert-curated clinical knowledge to guide GPT-4 reasoning at each decision node. GPT-4 generated binary decisions at each node to traverse the tree and classified MS diagnoses at terminal nodes. We evaluated performance against neurologist-assessed diagnoses and characterized hallucinations (non-factual, incongruent, irrelevant, over-reliant, and logical reasoning errors).

**Results.:** In this study cohort (mean age 40±13 years; 81% women) representative of the clinic population, GPT- 4 performed well in predicting MS diagnosis (84% accuracy, 79% precision, 74% recall, 91% specificity) using first neurology notes. Hallucinations occurred in 32 cases (26%), most commonly incoherence (75%) and overreliance (47%).

**Conclusion.:** A structured, LLM-guided decision framework can flag MS diagnoses from early clinical documentation. Large-scale studies are needed to mitigate hallucinations, validate this approach, and test implementation in clinical settings.

**HIGHLIGHTS:**

- Expert-curated MS knowledge guided GPT-4-based model through 2017 McDonald criteria
- Structured LLM framework identified MS in first neurology notes with 84% accuracy
- Binary decision trees made model reasoning more transparent and interpretable
- Hallucinations linked to incorrect MS classification; safeguards are needed
- Framework may generalize across complex criteria and reduce diagnostic delay

## 1 INTRODUCTION

Multiple sclerosis (**MS**) is a chronic inflammatory and neurodegenerative disorder of the central nervous system (**CNS**), affecting approximately 1 million people in the United States.^1,2^ Diagnosis relies primarily on the McDonald criteria, which require clinical, radiologic, and laboratory evidence; the updated 2024 McDonald criteria are not yet widely adopted in clinical practice.^3,4^ Despite improvements in diagnostic criteria, delays remain common, even in well-resourced healthcare systems.^5–7^ These delays arise across the diagnostic pathway. Patients must first recognize symptoms that are often nonspecific, such as visual disturbances, sensory changes, or gait instability. Access barriers may delay initial evaluation, with patients often first presenting in primary care or emergency settings, where clinicians must initiate further workup and specialist referral. Even for neurologists, MS mimics, the complexity of diagnostic criteria, and the lack of specific diagnostic biomarkers can further delay diagnosis. However, timely diagnosis of MS is crucial, as early initiation of disease-modifying therapy improves outcomes and reduces long-term disability.^8–10^

Large language models (**LLMs**) such as generative pre-trained transformers (**GPTs**) can synthesize information from clinic encounter notes, imaging reports, and laboratory data, and may flag complex diagnoses early. However, clinical deployment presents several challenges. LLMs designed for clinical reasoning tasks must demonstrate reliability, interpretability, and safety.^11,12^ Hallucinations may limit the accuracy and reliability, and the “black box” nature of LLMs limits interpretability.^13^ Although LLMs encode clinical knowledge, their domain- particular knowledge for specific diseases is variable.^14,15^ LLMs also demonstrate variable performance on complex medical tasks, including diagnosing MS and related disorders.^16–19^

In this proof-of-concept study, we examined the feasibility of flagging MS diagnoses from *the first neurology* clinical note using a structured, interpretable approach. We translated the 2017 McDonald criteria into a decision tree and guided GPT-4 through this framework using curated clinical knowledge. We evaluated GPT-4 performance against clinician-determined diagnoses and systematically characterized hallucinations to identify generalizable strengths and limitations of this approach.

## 2 METHODS

### 2.1 Data source and inclusion criteria

We randomly selected 125 patients (≥18 years) with at least one neurology clinic visit from an ongoing clinic cohort that began recruitment on January 2, 2017, including people with relapsing-remitting MS (**RRMS**), related neuroimmune conditions, and healthy controls. We excluded primary progressive MS (PPMS) because it has different diagnostic requirements in the 2017 McDonald criteria, and secondary progressive MS (SPMS) because it is not formally defined by diagnostic criteria but rather characterized by clinical course. Retrospective clinical records were accessed for research purposes on November 1, 2023. Authors with appropriate study authorization had access to information that could identify individual participants during data collection and chart review. We extracted the first neurology note, authored by either a general neurologist or MS subspecialist (**Fig. 1**), retaining the history of present illness, past medical history, review of systems, and diagnostic testing sections. We redacted the assessment and plan section and manually confirmed that no diagnosis information remained. We further manually redacted all protected health information except for dates (*e.g.,* encounter dates, symptom onset dates). Notes were input into a HIPAA-compliant Microsoft Azure instance of OpenAI GPT-4.

**Figure 1.**
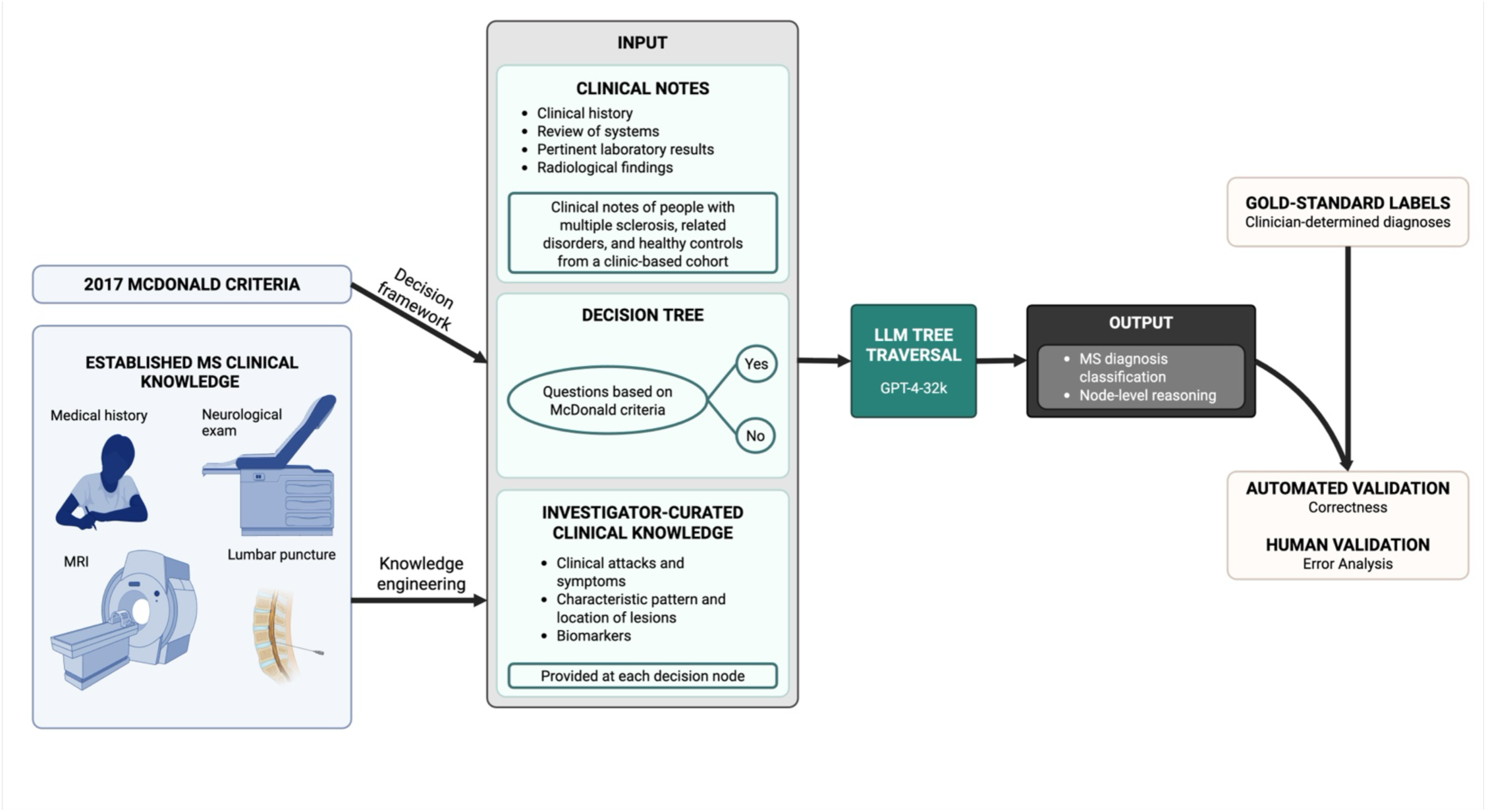
Study design. We developed a structured pipeline to identify multiple sclerosis (MS) diagnosis status from first neurology clinical notes without the neurologist’s assessment and plan. The 2017 McDonald criteria and an expert-curated MS knowledge base (including clinical history, neurologic examination, imaging, and cerebrospinal fluid findings) were used to construct a binary decision tree and provide domain-specific context. The knowledge base was iteratively refined based on model reasoning and reviewed by an MS specialist. Inputs included history, review of systems, laboratory results, and radiologic findings from first neurology notes of patients with MS, related disorders, and healthy controls in a clinic-based cohort. The large language model (GPT-4) was provided with the clinical note and curated MS knowledge and prompted to traverse the decision tree through sequential yes/no decisions. At terminal nodes, the model generated a classification of MS diagnosis status. Outputs were compared with clinician-determined diagnoses (gold-standard labels) obtained through chart review and confirmed by an MS specialist; performance was assessed using automated metrics, and errors were further characterized through human review of hallucinations.

### 2.2 Ethics approval

The University of Pittsburgh Institutional Review Board approved this study (STUDY19080007). All participants provided written informed consent allowing access to their medical records.

### 2.3 McDonald criteria

We completed analyses when the 2017 McDonald criteria were still the latest MS diagnostic criteria. The 2017 McDonald criteria demonstrate high sensitivity for MS detection, but are not intended to differentiate MS from related disorders.^3,20^ The criteria can be applied in a person with an attack or relapse at onset (*i.e.,* the occurrence of a new or recurrent neurological symptom lasting over 24 hours, with or without recovery, in the absence of fever or infection). The attack should be confirmed by objective clinical evidence such as abnormal physical examination findings or diagnostic testing results that correspond to the anatomical location suggested by the attack symptoms. The criteria further require objective clinical evidence or reasonable historical evidence of demyelination in at least two different locations in the CNS (*i.e.,* disseminated in space **[DIS]**) or at different points in time (*i.e.,* disseminated in time **[DIT]**). DIS is based on either evidence of an additional clinical attack implicating a different region of the CNS than the first attack, or symptomatic or asymptomatic lesion presence in two distinct CNS locations (periventricular, cortical / juxtacortical, infratentorial, or spinal cord). DIT is based on either an additional clinical attack occurring at least 30 days after the first attack, simultaneous presence of both enhancing and non-enhancing symptomatic or asymptomatic lesions in MS-typical CNS locations, a new or enhancing lesion compared to baseline, or the presence of oligoclonal bands in cerebrospinal fluid (**CSF**).

### 2.4 Decision tree

We translated the 2017 McDonald criteria into a binary decision tree to enable structured, stepwise evaluation (**Fig. 2**).^3,12^ First, the algorithm identifies alternative diagnoses by searching for objective clinical evidence of alternative neuroimmune disorders (*e.g.,* neuromyelitis optica [**NMO**], myelin oligodendrocyte glycoprotein antibody-associated disease [**MOGAD]**), as the MS criteria do not apply in these cases. It then evaluates whether there is objective evidence of a clinical attack at onset, followed by assessment of DIS and DIT criteria. Terminal nodes classify MS diagnosis status as MS confirmed, MS not confirmed (lacking evidence of DIS and DIT), MS not confirmed (lacking evidence of DIS), MS not confirmed (lacking evidence of DIT), or MS not confirmed (alternative diagnosis).

**Figure 2.**
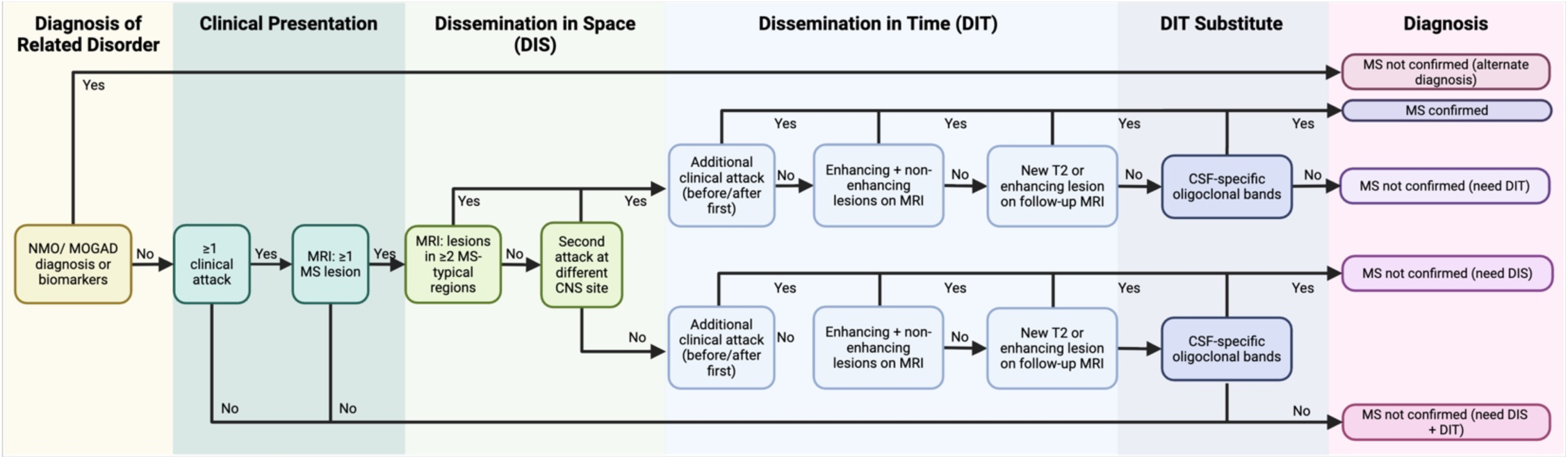
2017 McDonald criteria decision tree. A decision tree is a flowchart-like tool used to guide clinical decision-making through a sequence of binary (yes/no) questions that ultimately lead to a diagnostic classification. We manually constructed a decision tree representing the 2017 McDonald criteria for MS diagnosis, which first considers clinical presentation and then incorporates additional data (e.g., MRI and cerebrospinal fluid findings) to determine dissemination in space (DIS) and dissemination in time (DIT). We provided the large language model (GPT-4) with expert-curated clinical knowledge (e.g., clinical attacks, central nervous system lesions, and biomarkers), developed from established diagnostic criteria and reviewed by MS clinical experts. The decision tree guided the LLM through the diagnostic process: at each node, the model answered the node question and provided a rationale. At the final node, the LLM classified diagnosis status as MS confirmed or MS not confirmed (alternate diagnosis, lacking evidence of DIS and DIT, DIS only, or DIT only).

### 2.5 LLM

During analyses, OpenAI GPT-4 was a readily available LLM. We leveraged a HIPAA-compliant Microsoft Azure instance of GPT-4-32k and set the seed (9631) for reproducibility. GPT-4 was used for node-level reasoning within the decision tree to derive MS diagnosis status. Two clinical domain experts (SV, MD) curated clinical knowledge based on the 2017 McDonald criteria (*e.g.,* clinical attacks, lesion characteristics, biomarkers). The clinical knowledge was iteratively refined through systematic evaluation of model reasoning, with modifications made to reduce misinterpretation and improve alignment with the 2017 McDonald criteria. The curated clinical knowledge was provided to GPT-4 alongside the clinical note at each decision node (**Supplementary Methods**). GPT-4 generated a binary (yes/no) response at each node to guide traversal of the decision tree and provided a rationale for its decision. We used a low temperature (0.2) during node traversal to promote deterministic outputs, with a top-p parameter of 0.6 to limit diversity in reasoning. At terminal nodes, we used a temperature of 0.5 to balance coherence and flexibility for classification of MS diagnosis status.

### 2.6 GPT evaluation

We compared GPT-4–predicted diagnoses with clinician-determined gold-standard labels, extracted from the assessment and plan of the neurology note, and confirmed by an MS subspecialist (ZX). Review was conducted post hoc, without feedback to the model during traversal or diagnosis. Model performance was evaluated using standard metrics, including accuracy, precision, recall, and specificity. We further characterized hallucinations in the incorrect responses. While hallucinations are broadly categorized as factuality (response contradicts established knowledge) or faithfulness (response deviates from provided input), we extended these categories using a dialogue-level hallucination evaluation framework to assess errors at each decision node response.^11,13,21^ Since GPT-4 generated a rationale at each decision node, this approach enabled evaluation of hallucinations at the node level rather than at the sentence or full-response level.^21^ Hallucinations were classified as non-factual (output contradicts facts), incoherent (output contradicts input context or dialogue is self-contradictory), irrelevant (output is unrelated to the topic), overreliant (excessive reliance on provided context without applying provided knowledge), or logical reasoning errors (error in logic or understanding). Responses could exhibit more than one hallucination type.

### 2.7 Code availability

Analysis code is publicly available on GitHub.^22^

### 2.8 Data availability

Anonymous summary-level data will be publicly available.^22^ Patient-level data will not be publicly available because patient-level clinical data, whether de-identified or containing limited protected health information such as dates of clinical events, or even if anonymous due to the potential risk of re-identification, are universally subject to the rules and regulations of the healthcare system, which is affiliated with but not the same as the primary academic institution of the study investigators.

## 3 RESULTS

### 3.1 Patient characteristics

The study comprised 125 patients with a mean age of 40±12 years at first neurology note (**Table 1**): 105 with MS (38±12 years), 10 with other neuroimmune disorders (44±16 years), and 10 healthy controls (48±11 years). Patients were predominantly women (81%), White (90%), and non-Hispanic (99%), with a low mean neighborhood-level deprivation (area deprivation index 0.31±0.11). Among people with MS, the delay between neurological symptom onset and the first documented neurology clinic visit was substantial (diagnostic delay mean±SD=3.37±6.40 years).

**Table 1.** Cohort characteristics.

| Characteristic | Overall | RRMS | Related Disorder | Healthy Control |
| --- | --- | --- | --- | --- |
| Number of patients | 125 | 105 | 10 | 10 |
| Women, N (%) | 101 (81%) | 88 (84%) | 5 (50%) | 8 (80%) |
| Race, N (%) |  |  |  |  |
| Black | 10 (8%) | 8 (8%) | 2 (20%) | 0 (0%) |
| Asian | 2 (2%) | 2 (2%) | 0 (0%) | 0 (0%) |
| White | 113 (90%) | 95 (90%) | 8 (80%) | 10 (100%) |
| Non-Hispanic, N (%) | 124 (99%) | 105 (100%) | 9 (90%) | 10 (100%) |
| Area deprivation index, mean (SD) | 0.31 (0.11) | 0.31 (0.11) | 0.31 (0.11) | 0.27 (0.07) |
| Age at first neurology note, mean (SD) | 39.52 (12.41) | 38.37 (11.88) | 43.50 (15.99) | 47.60 (11.30) |
| Age at neurological symptom onset, mean (SD) |  | 35 (10.70) |  |  |
| Diagnostic delay, mean (SD) |  | 3.37 (6.40) |  |  |
| Disease-modifying therapy class at the first neurology note, N (%) |  |  |  |  |
| Interferon-beta |  | 8 (8%) |  |  |
| Glatiramer acetate |  | 13 (12%) |  |  |
| Fumarate |  | 16 (15%) |  |  |
| Alpha4-integrin blocker |  | 19 (18%) |  |  |
| B cell depletion therapy |  | 28 (27%) |  |  |
| None |  | 21 (20%) |  |  |

We report baseline characteristics at the first neurology note for the overall cohort and stratified by the diagnostic categories assigned in the clinic-based cohort. Demographic characteristics are reported for the overall cohort, while MS-specific clinical factors (age at neurological symptom onset, diagnostic delay, and disease-modifying therapy class at the first neurology note) are reported for MS patients only. Area deprivation index values range from 0 to 1, with higher values indicating greater neighborhood-level socioeconomic disadvantage. Continuous variables are reported as mean (SD) and categorical variables as N (%).

Based on the neurologist’s assessment and plan in the first neurology note, 66 of 125 patients (53%) had a confirmed MS diagnosis (**Table 2**). Of the remaining 59 patients, 34 (27%) lacked evidence of both DIS and DIT, 12 (10%) lacked DIS only, 8 (6%) lacked DIT only, and 5 (4%) had alternative diagnoses. Of the 125 neurology notes analyzed with GPT-4, 32 (26%) were authored by general neurologists and 93 (74%) by MS specialists.

**Table 2.** Performance of MS diagnosis classification.

| **MS Diagnosis Classification** | **N** | **Accuracy** | **Precision** | **Recall** | **Specificity** | **F1 score** |
| --- | --- | --- | --- | --- | --- | --- |
| Overall cohort | 125 | 0.84 [0.77, 0.90] | 0.79 [0.62, 0.90] | 0.74 [0.56, 0.87] | 0.91 [0.82, 0.96] | 0.75 |
| MS confirmed | 66 | 0.79 [0.71, 0.86] | 0.90 [0.78, 0.97] | 0.68 [0.56, 0.79] | 0.92 [0.81, 0.97] | 0.78 |
| MS not confirmed (lacking evidence of DIS and DIT) | 34 | 0.86 [0.79, 0.92] | 0.71 [0.55, 0.84] | 0.85 [0.69, 0.95] | 0.87 [0.78, 0.93] | 0.77 |
| MS not confirmed (lacking evidence of DIS) | 12 | 0.89 [0.82, 0.94] | 0.44 [0.22, 0.69] | 0.67 [0.35, 0.90] | 0.91 [0.84, 0.96] | 0.53 |
| MS not confirmed (lacking evidence of DIT) | 8 | 0.94 [0.89, 0.98] | 0.55 [0.23, 0.83] | 0.75 [0.35, 0.97] | 0.96 [0.90, 0.99] | 0.63 |
| MS not confirmed (alternative diagnosis) | 5 | 1.00 [0.97, 1.00] | 1.00 [0.48, 1.00] | 1.00 [0.48, 1.00] | 1.00 [0.97, 1.00] | 1.00 |

We report performance metrics for MS diagnosis status classification with 95% confidence intervals. Metrics were computed by comparing GPT-4–predicted classifications with clinician-determined gold-standard MS diagnosis status labels extracted from the assessment and plan section of the first neurology note. Gold-standard labels were redacted from the input provided to GPT-4 and independently verified by an MS subspecialist. For the overall cohort, we report the weighted average of each metric across all classes. *DIS*, dissemination in space; *DIT*, dissemination in time

### 3.2 GPT performance

GPT-4 demonstrated overall robust predictive performance with an average accuracy of 84%, precision of 79%, recall of 74%, and specificity of 91% across all classes (**Table 2**). For confirmed MS, GPT-4 demonstrated high precision (90%) but comparatively lower recall (68%), suggesting that some true MS patients were missed, potentially those with atypical presentations or early-stage disease. While GPT-4 demonstrated robust performance in detecting alternative diagnoses (100% specificity and precision), it achieved high specificity (87– 96%) but comparatively low precision (44–71%) in recognizing patients lacking evidence of DIS, DIT, or both. Of the 125 first neurology notes, hallucinations were detected in GPT-4 responses for 32 (26%) notes. Incoherence hallucinations were the most common (75%), followed by overreliance (47%), irrelevance (19%), reasoning (19%), and non-factual (16%) hallucinations (**Table 3**). Hallucinations were typically associated with inaccurate diagnosis classification. Upon detailed review, several themes emerged. Incoherence hallucinations often occurred when GPT-4 inconsistently applied assumptions provided to the model or failed to fully interpret information within the clinical note. For example, GPT-4 could correctly identify the presence of enhancing lesions, but failed to detect non-enhancing lesions, despite the clinical context stating that lesions should be assumed to be non-enhancing by default (**Supplementary Methods, S-Table 1**). We also observed that GPT- 4 could correctly identify the presence of a clinical attack, but erroneously stated that the neurological symptoms were not related to MS or did not last longer than 24 hours. Overreliance hallucinations stemmed from conservative reasoning due to excessive dependence on the clinical context pertaining to magnetic resonance imaging (**MRI**). In one example, GPT-4 correctly identified the presence of a periventricular lesion, but failed to categorize this lesion as being present in an MS-typical CNS region. In another instance, GPT-4 produced an overreliance hallucination by failing to recognize punctate lesions, signal anomalies, or changes consistent with demyelinating disease as lesions. Irrelevance hallucinations typically contained information from the patient note or clinical context, and were less likely to be associated with incorrect diagnosis classification. Reasoning hallucinations predominantly stemmed from incorrect calculation of the duration between dates mentioned in the clinical note. This resulted in incorrect DIS and DIT identification, as GPT-4 could not correctly interpret the time elapsed between distinct clinical attacks or lesion occurrences. Non-factual errors typically stemmed from imperfect understanding of neurological symptoms. For example, GPT-4 failed to identify atrial lesions as periventricular lesions, cerebellar lesions as infratentorial lesions, and gray–white junction lesions as juxtacortical lesions. This led GPT-4 to inaccurately state that there were no lesions in MS-typical CNS regions.

**Table 3.** Frequency of hallucinations by category.

| Hallucination Category | Hallucinations, n (% of N=32) | Hallucinations Resulting in Inaccurate Diagnosis Classification, n (% of N=32) |
| --- | --- | --- |
| Non-factual | 5 (16) | 4 (13) |
| Incoherent | 24 (75) | 21 (66) |
| Irrelevant | 6 (19) | 1 (3) |
| Overreliant | 15 (47) | 15 (47) |
| Reasoning | 6 (19) | 6 (19) |

Hallucinations were identified through manual chart review of GPT-4 responses at each decision node using a predefined classification framework. Values represent the number and percentage of the 32 cases in which at least one hallucination type was detected. Categories were not mutually exclusive. The number of hallucinations associated with inaccurate diagnostic classification are also reported. Hallucination categories include non-factual (output contradicts facts), incoherent (output contradicts input context or dialogue is self-contradictory), irrelevant (output is unrelated to the topic), overreliant (excessive reliance on provided context without applying provided knowledge), or logical reasoning errors (error in logic or understanding).

## 4 DISCUSSION

Our study demonstrates a framework for LLM-guided, interpretable decision trees, augmented by disease-specific clinical knowledge, for deconstructing complex diagnostic criteria and identifying a challenging neurological condition from initial neurology clinical documentation. Using MS and the McDonald criteria to illustrate the framework, we first converted a complex clinical algorithm of disease diagnosis into a computable form using a binary decision tree, and curated domain-specific knowledge with iterative revisions based on model reasoning. We then prompted an LLM (GPT-4) at each decision node with specific clinical questions to obtain contextually relevant outputs while maintaining interpretability. While GPT-4 accurately predicted MS diagnosis status blinded to the neurologist’s assessment in most patients, the risk of hallucination remained. Recognizing and mitigating hallucination in LLM-enhanced diagnostic support systems will be critical for future clinical implementation. Broadly, this study serves as a proof-of-concept case study and offers a potentially generalizable framework for integrating structured clinical reasoning with LLMs to support earlier diagnosis of otherwise challenging diseases with complex criteria, while highlighting the methodological safeguards critical to the clinical deployment of LLM-enhanced diagnostic support systems.

### 4.1 LLM-enhanced diagnostic support systems could reduce diagnostic delay

Using this approach to flag MS diagnoses from the first neurology clinic documentation by detecting patterns consistent with an MS diagnosis could potentially reduce one of the many roadblocks in the MS diagnostic delay. There are other roadblocks between initial neurological symptom onset and the first neurology visit. This study used the first neurology documentation, capturing patients with neurological symptoms who were referred to general neurologists or MS specialists within a large healthcare system. However, our approach may not be generalizable to initial evaluations in primary care, urgent care, emergency department, or inpatient settings. Future studies should evaluate the approach in other healthcare settings.

Along the MS diagnostic pathway (*i.e.,* from initial evaluation in a primary care or emergency room setting, followed by evaluation by a general neurologist, and potentially an MS specialist), implementing an LLM-enhanced diagnostic support system tailored to complex neurological disorders such as MS at the point of care, particularly in general neurology settings, could be beneficial. Access to MS specialists is limited even in large urban areas. Although general neurologists may be familiar with the MS diagnostic criteria, they are still likely to refer patients to MS specialists due to diagnostic uncertainty. Implementing an LLM-enhanced diagnostic support system for MS in this setting could accelerate diagnostic testing and enable early treatment initiation, which can improve patient outcomes and equitable care across diverse patient populations.

### 4.2 Challenges in developing LLM-enhanced diagnostic support systems for MS diagnosis

Our proof-of-concept study highlights the need for rigorous validation of LLM performance and hallucination mitigation to ensure the accuracy, reliability, and interpretability of LLM-enhanced diagnostic support systems before clinical implementation, thereby building trust among clinicians and patients.^23–26^

Prior studies applying natural language processing and LLM-based approaches for clinical tasks in neurology and MS demonstrate variable performance, likely due to task structuring and complexity in addition to the underlying models. While the models generally perform well on well-defined tasks such as MRI report classification^27^ and structured knowledge assessments,^15^ their performance declines in open-ended diagnostic reasoning involving real-world clinical cases.^28,29^ Structured diagnostic frameworks, such as the decision tree-guided approach implemented in this study, can constrain model reasoning and improve reliability.

Hallucination mitigation for LLMs remains a major challenge even with model improvement.^30^ Although several hallucination mitigation strategies exist, no single strategy is sufficient, and effective mitigation requires layered safeguards.^30,31^ Retrieval-augmented generation supplements the model’s internal knowledge with relevant external documents at inference time to produce more accurate, contextually grounded outputs.^32,33^ Prompt engineering strategies reduce hallucinations by constraining model reasoning.^34^ For example, chain-of-thought prompting may improve performance by guiding the model through intermediate reasoning steps before arriving at a final answer. Other hallucination mitigation strategies include context-aware decoding to prioritize the provided clinical context over the model’s inherent knowledge,^35^ human-in-the-loop systems in which clinician feedback refines model behavior,^36^ uncertainty quantification to flag low-confidence outputs for clinician review,^37,38^ and emerging multi-agent frameworks in which specialized LLM agents cross-validate outputs.^39^

LLM interpretability—understanding how these models generate outputs and making their internal decision-making processes more transparent—remains a major challenge. Although we prompted GPT-4 to provide its rationale at each decision node, we were unable to determine the extent to which GPT-4 relied on the clinical note, curated knowledge, or inherent training data in its responses. Strategies to improve LLM interpretability, including the use of stepwise reasoning, chain-of-thought prompting, explanations from retrieval-augmented generation, and integration of explainable artificial intelligence techniques to improve transparency and interpretability, are an active area of development.^40^

### 4.3 Strengths and Limitations

The study has several strengths. First, the approach flagged MS diagnosis from the first neurology note, blinded to the neurologist’s assessment and plan, often preceding a formal diagnosis. This proof-of-concept study highlights the potential for LLM-enhanced tools to reduce diagnostic delay at the point of care. Second, we integrated a transparent, rule-based decision tree for structured reasoning with LLM-based reasoning using curated clinical knowledge, which can improve accuracy and interpretability while maintaining flexibility.^41,42^ Reasoning can be constrained by decision trees to match clinical algorithms and mitigate hallucination risk. LLMs enable synthesis of complex information across clinical notes and the knowledge base. Third, we validated LLM-predicted MS diagnosis status using gold-standard labels, systematically characterized the hallucinations produced, and identified pitfalls of LLM-enhanced diagnostic support systems that need to be addressed before clinical implementation. Moreover, we do not view the use of an older version of McDonald criteria or an older LLM as limitations for this proof-of-concept study. The 2024 McDonald criteria are not yet widely adopted and are not yet consistently reflected in clinical documentation. The newer LLMs may offer incremental improvement over the model used in this study, but the overall framework is disease- and model-agnostic, and potentially generalizable to other complex conditions.

There are also limitations given the proof-of-concept intention. First, we applied several exclusion criteria. To simplify the LLM-guided decision trees, we excluded PPMS because of its different diagnostic requirements in the 2017 McDonald criteria. We excluded SPMS because it is better characterized by clinical course rather than diagnostic criteria. We also excluded other rare neuroimmune disorders (except for NMO and MOGAD as disease controls). Second, the cost of running the LLM largely limited the sample size. In this study population randomly selected from a large clinic cohort, the initial neurology documentation included notes from both general neurologists and MS subspecialists, and patients came from a large catchment area spanning urban, suburban, and rural settings. Third, model performance may vary with documentation practices and quality, but we were not able to carefully examine this question. Finally, we built a curated knowledge base using established diagnostic criteria and expert review, with iterative refinement informed by evaluation of model reasoning. Although this approach improves clinical alignment, the resulting knowledge base remains static at inference and is not dynamically updated, which may limit scalability as clinical knowledge evolves. Retrieval-augmented generation could address this limitation by enabling incorporation of up-to-date medical information.

### 4.4 Future directions

Future directions of this work include expanding sample size, performing external validation across healthcare systems, and implementing hallucination mitigation strategies. While applying the 2024 McDonald criteria may improve diagnostic performance over the 2017 McDonald criteria, it also raises considerations for generalizability and implementation, particularly in settings without access to specialized imaging or laboratory tests.^4^ Our goal is not to establish a definitive diagnostic standard, but to demonstrate a generalizable framework for translating complex clinical criteria into computable, LLM-enhanced decision processes that can be adapted as standards evolve. Accordingly, the framework will need to be iteratively updated alongside changes in diagnostic criteria and clinical practice. Prospective evaluation, ideally through a randomized controlled trial, will be essential to determine whether LLM-enhanced diagnostic support reduces diagnostic delay while maintaining clinician trust and patient safety.

## 5 CONCLUSION

Our study demonstrates a potential road map for integrating structured clinical reasoning with LLMs to support the diagnosis of otherwise challenging diseases with complex diagnostic criteria such as MS. We anticipate that the accuracy, reliability, and interpretability of LLM-enhanced diagnostic support systems will improve as LLMs evolve. However, while LLM-enhanced diagnostic support systems may have the potential to reduce the diagnostic delay of complex conditions, iterative refinement guided by clinician feedback and rigorous validation in real-world settings before clinical deployment will be crucial. Ultimately, future work should evaluate how LLMs work alongside clinicians to supplement, not replace, clinician expertise.

## ACKNOWLEDGEMENTS

This work was supported by the National Institutes of Health under award R01NS098023, the University of Pittsburgh Clinical Scientist Training Program, and the University of Pittsburgh Clinical and Translational Science Institute under NIH award UL1TR001857. We thank Dr. Judy Chang and Dr. Scott Rothenberger for their thoughtful feedback and suggestions on this manuscript.

## Declaration of Conflicts of Interest

The authors declared no potential conflicts of interest with respect to the research, authorship, and/or publication of this article.

## Notes

### Competing Interest Statement

The authors have declared no competing interest.

