## Supplementary Materials for "Large Language Model - Enhanced Decision Tree Framework for Identifying Multiple Sclerosis Diagnoses from Clinical Documentation"

### SUPPLEMENTARY MATERIAL

**S-Method 1.** Expert-curated clinical knowledge provided at each decision node.

**S-Method 2.** Model development.

**S-Table 1.** Examples of GPT-4 hallucinations during decision tree traversal.

**S-Figure 1.** Example of node-level model reasoning for clinical attack determination.

**S-Figure 2.** Illustrative example of node-level model reasoning with incoherent and irrelevant hallucinations.

**S-Figure 3.** Comparison of Model Configurations for MS Diagnosis Derivation.

#### SUPPLEMENTARY METHODS

#### S-Method 1. Expert-curated clinical knowledge provided at each decision node.

Clinical knowledge was curated by the study team based on the 2017 McDonald criteria and supporting clinical and radiologic literature. Content was iteratively refined through expert review and evaluation of model reasoning to ensure alignment with diagnostic criteria and reduce misinterpretation. The content summarizes the node-specific clinical context provided to the model.

**Diagnosis of Related Disorder Node 1: Does the patient have a diagnosis of NMO or MOGAD, or positive laboratory findings of NMO-IgG, AQP4-Ab, or MOG-Ab?**

Certain neurological conditions with similar clinical presentations to multiple sclerosis (MS) can be differentiated from MS by findings of specific serological markers. These include neuromyelitis optica (NMO) and myelin oligodendrocyte glycoprotein antibody-associated disease (MOGAD). Laboratory evidence of aquaporin-4 antibodies (AQP4-IgG, or NMO-IgG) in the serum or myelin oligodendrocyte glycoprotein (MOG) antibodies in the serum or cerebrospinal fluid are not present in MS and represent these distinct diseases. Patients with diagnoses of NMO or MOGAD may be seropositive or seronegative on antibody testing. Patients with a diagnosis of NMO or MOGAD or any laboratory evidence of the specific biomarkers for NMO or MOGAD do not have a diagnosis of MS. These patients should be categorized as having an alternative diagnosis that is distinct from MS.

**Clinical Presentation Node 1: Does the patient have at least one clinical attack?**

\# *clinical attack*

A clinical attack (also known as a relapse or exacerbation or flare) is characterized by the occurrence of continuous new or worsening neurological symptoms. Clinical attacks can include patient-reported symptoms or any objective findings reflective of a monofocal or multifocal inflammatory demyelinating event within the central nervous system. Rather than counting individual symptoms as discrete clinical attacks, only cohesive periods of continuous neurological symptoms should be identified as clinical attacks. This is because several diverse neurological symptoms may constitute a single clinical attack.

In order to be considered as a true clinical attack, the following criteria need to be met. First, neurological symptoms should occur for at least 24 hours with or without recovery. Second, neurological symptoms must occur in the absence of fever, infection, stress (including an adverse psychosocial event) or heat exposure (including warm weather, fever, or exercise). Third, discrete clinical attacks should be at least 30 days apart, i.e., there must be at least a 30-day period without any neurological symptoms between clinical attacks. If neurological symptoms manifest within the 30-day interval, the symptom should be considered to be a part of the previous clinical attack (unless explicitly stated by the clinician, though this is rare). Only count confirmed clinical attacks that meet the aforementioned criteria. Do not count suspected clinical attacks.

If patients are noted as having a monophasic demyelinating disorder, this means that they have had one previous clinical attack. The neurological symptoms associated with the monophasic demyelinating episode count as the first clinical attack.

Mild fleeting symptoms, such as muscle spasms, pain, or abnormal sensations, may frequently occur for a few minutes to hours in people with multiple sclerosis (MS). These do not meet the criteria for clinical attack and should not be counted as discrete clinical attacks. Likewise, non-specific symptoms such as fatigue, depression, and vague cognitive complaints such as “brain fog” cannot be localized neuroanatomically and should not be considered as clinical attacks. If a symptom has been constant (i.e., never went away from onset) and has worsened in severity, it is not typically considered an attack.

The date of the patient note should be used as a reference point for the timing of a clinical attack. For example, if a patient note was written on 12/30/2015, and the note refers to symptoms starting “3 weeks ago”, then all symptoms occurred in the three-week period between 12/9/2015 and 12/30/2015.

\# *typical MS presentations*

Clinical attacks are characterized by continuous neurological symptoms that involve any of the following central nervous system regions: cranial nerves, brainstem, cerebellum, cerebrum, or spinal cord.

The most common presentations of clinical attacks involving the brainstem and cranial nerves are optic neuritis, visual loss, internuclear ophthalmoplegia, facial sensory loss, hearing loss, dysarthria, and diplopia (e.g., due to abducens nerve palsy). Any time these symptoms occur, you should assume they are clinical attacks, unless the symptoms are noted to occur for less than 24 hours or intermittently.

The most common presentations of clinical attacks involving the cerebellum are ataxia, balance problems, gait disturbance, and oscillopsia. Any time these symptoms occur, you should assume they are clinical attacks, unless the symptoms are noted to occur for less than 24 hours or intermittently.

The most common presentations of clinical attacks involving the cerebrum and spinal cord are limb sensory loss, upper motor neuron signs, acute transverse myelopathy, and sensations of a tight band around the trunk or abdomen (also called a “MS hug”). Any time these symptoms occur, you should assume they are clinical attacks, unless the symptoms are noted to occur for less than 24 hours or intermittently.

Some symptoms are commonly observed in MS, but are not specific indicators of disease. These include spasticity, paresthesia (numbness or tingling), pain, weakness, vertigo, and dizziness. Any time these symptoms occur, you should assume they are not clinical attacks, unless the symptoms are noted to occur persistently for over 24 hours.

Certain symptoms occur in MS that should not be counted as clinical attacks, such as urinary incontinence (spastic bladder), Lhermitte sign (an electrical sensation along the back and/or limbs elicited with neck flexion), headache, alteration of consciousness, trigeminal neuralgia, fatigue, depression, Uhthoff phenomenon (exacerbation of symptoms due to heat, warm weather, or exercise), and cognitive decline or cognitive impairment.

You should be very conservative with counting clinical attacks based on the above criteria. Do not count clinical attacks that fall outside of these guidelines.

\# *pseudo-exacerbations*

Fever, infection, stress (including an adverse psychosocial event), or heat exposure (including warm weather, fever, or exercise) can trigger pseudo-exacerbations among people with MS. Pseudo-exacerbations are not considered clinical attacks.

**Clinical Presentation Node 2: Does the patient have MRI evidence of at least one MS lesion?**

\# *MRI lesions*

MRI is the most sensitive method to detect monofocal and multifocal demyelinating lesions in multiple sclerosis (MS). Demyelinating lesions must be associated with a specific region of the central nervous system (brain or spinal cord) and detected on MRI.

On MRI, both gadolinium-enhancing and non-enhancing lesions may be associated with MS. Gadolinium-enhancing lesions and new or enlarging active lesions are indicative of an ongoing clinical attack. Gadolinium-enhancing lesions reflect acute inflammation at the time of imaging whereas active T2 lesions reflect a previous focal inflammatory lesion that has developed since the prior MRI, but the exact time point of this activity cannot be identified. For example, a description may mention "MRI showed enhancement of the optic nerve" when there is enhancing lesion in the optic nerve.

Inactive lesions (also known as non-enhancing lesions or T2 hyperintense lesions) are indicative of a prior clinical attack or old regions of disease activity. Lesions should be assumed to be non-enhancing unless explicitly stated to be enhancing.

Do not include superficial or nonspecific lesions as they are not associated with any brain region. For example, “few nonspecific punctate superficial white matter abnormalities were seen on T2 and FLAIR sequences” would not count as a MS lesion. This is because these are superficial, nonspecific signals not associated with any brain region. As the observed abnormalities cannot be localized to a specific anatomical area or structure within the brain, we would not be able to verify that these were MS lesions.

\# *MRI settings*

Lesions appear differently on MRI depending on the MRI imaging settings, lesion location, and whether the lesion is part of a current or previous clinical attack.

T1-weighted MRI: MS lesions are typically isointense to hypointense (T1 black holes). Callososeptal interface may have multiple small hypointense lesions or the corpus callosum may appear thinned. In T1-weighted MRI with gadolinium-based contrast, active MS lesions show enhancement, whereas inactive lesions from previous attacks do not show enhancement.

T2-weighted MRI: MS lesions are typically hyperintense, and acute lesions often have surrounding edema.

Fluid-attenuated inversion recovery (FLAIR): MS lesions are typically hyperintense.

**Dissemination in Space Node 1: Does the patient have MRI evidence of at least one MS lesion in two distinct MS typical regions (periventricular, cortical or juxtacortical, infratentorial, or spinal cord)?**

We provided the context given for the question, **“Does the patient have MRI evidence of at least one MS lesion?”** and additional context as noted below.

*# lesions in at least two MS-typical regions*

This question asks whether the description mentions the presence of T2 hyperintense lesions in at least two of the multiple sclerosis (MS) typical regions. MS typical regions include the following areas of the central nervous system: periventricular, cortical or juxtacortical, infratentorial, and the spinal cord. Lesions in the deep white matter, subcortical brain regions, and optic nerve should not be counted as lesions in MS typical regions or towards the McDonald criteria for dissemination in space. Lesions in other areas or nonspecific locations should not be counted toward any of the MS typical regions.

Periventricular lesions are T2 hyperintense lesions that abut the lateral ventricles without white matter in between, including lesions in the corpus callosum but excluding lesions in deep gray matter structures. Pericallosal lesions are considered as periventricular lesions as they are located near the lateral ventricles and the corpus callosum. If there are pericallosal lesions, callosal lesions, or lesions in the corpus callosum, they should be counted as periventricular lesions and as one region towards the McDonald criteria for dissemination in space.

Cortical lesions are located within the cerebral cortex. Typically, special MRI techniques such as double inversion recovery, phase-sensitive inversion recovery, and magnetization-prepared rapid acquisition with gradient echo sequences are required to visualize these lesions. Juxtacortical lesions are T2-hyperintense lesions that abut the cortex and are not separated from it by white matter.

Infratentorial lesions are T2-hyperintense lesions in the brainstem (typically near the surface), cerebellar peduncles, or cerebellum.

Spinal cord lesions are T2 hyperintense lesions that occur in the cervical, thoracic, or lumbar spinal cord seen on T2 plus short tau inversion recovery, proton-density images, or other appropriate sequences, or in two planes on T2 images.

**Dissemination in Space Node 2: Does this patient have an additional clinical attack involving a different central nervous system location than the first clinical attack?**

We provided the context given for the question, **“Does the patient have at least one clinical attack?”** and additional context as noted below.

\# *criteria for a clinical attack at another site*

An additional clinical attack is defined as a clinical attack that occurs at a distinct site within the central nervous system in comparison to the previous clinical attack. Typically, it involves different neurological symptoms (or a different location) than the previous clinical attack. For attacks to be at two distinct locations, each attack should localize to a different part of the brain, spinal cord, or to the optic nerve.

Here are a few examples that illustrate the concept of clinical attacks involving different central nervous system sites:

If the first clinical attack involves right arm weakness and the second clinical attack involves left arm weakness, this would be considered an additional clinical attack at another site.

If the first clinical attack involves bilateral paresthesia and weakness in the arms and the second clinical attack involves bilateral leg weakness, this would be considered an additional clinical attack at another site.

If the first clinical attack involves diplopia and the second clinical attack involves unilateral hearing loss, this would be considered an additional clinical attack at another site.

If the first clinical attack involves diplopia and the second clinical attack involves ataxia, this would be considered an additional clinical attack at another site.

If the first clinical attack involves optic neuritis and the second clinical attack involves ataxia, this would be considered an additional clinical attack at another site.

**Dissemination in Time Node 1: Does the patient have two confirmed clinical attacks occurring at least 30 days apart?**

We provided the context given for the question, **“Does the patient have at least one clinical attack?”**

**Dissemination in Time Node 2: Are enhancing lesions and non-enhancing lesions present together on MRI at any time?**

We provided the context given for the question, **“Does the patient have MRI evidence of at least one MS lesion?”** pertaining to MRI lesions.

**Dissemination in Time Node 3: Is there evidence of a new hyperintense T2 lesion or new enhancing lesion on follow-up or repeat MRI?**

We provided the context given for the question, **“Are enhancing lesions and non-enhancing lesions present together on MRI at any time?”** and additional context as noted below.

A radiological attack is defined as either the presence of any new T1 gadolinium-enhancing lesion, or any new or enlarging T2-FLAIR lesion in comparison to the prior scan. Any lesion that is new compared to a previous MRI from the same area of the central nervous system should be counted as a new lesion. For example, new lesions in the frontal cortical region on a repeat brain MRI compared to a previous cranial MRI should be counted as a new lesion. Do not compare MRIs from different areas of the body. For example, do not compare a brain MRI to a spinal cord MRI when assessing new lesions.

**Dissemination in Time Substitute Node 1: Is there evidence of cerebrospinal fluid (CSF)-specific oligoclonal bands?**

No context was provided.

#### S-Method 2. Model development.

To inform model development, we compared alternative ways of structuring the task of deriving MS diagnosis status from a clinical note using the same LLM with varying degrees of clinical guidance. We evaluated five configurations: (1) clinical note alone with direct diagnosis generation; (2) clinical note with expert-curated clinical knowledge and a narrative description of the 2017 McDonald criteria; (3) the decision tree based on the 2017 McDonald criteria alone; (4) the decision tree with expert-curated clinical knowledge (**primary approach**); and (5) the decision tree with expert-curated clinical knowledge and examples for few-shot learning (**S-Figure 3**).

Each configuration was applied to a subset of three clinically complex notes to assess effects on model reasoning and diagnostic classification. Model performance was evaluated using gold-standard labels, and the best-performing configuration was selected for the primary analysis. An illustrative example of the clinical note excerpt, reference standard, node-level prompts, and configuration outputs are provided below.

##### Clinical note excerpt

**DATE OF VISIT:** Late 2008.

**REASON FOR VISIT:** Unsteady gait with right-sided weakness.

**HISTORY OF PRESENT ILLNESS:** This is a female [in her 50s] who was seen in our clinic remotely back in 2004. She was referred to Dr. [NAME] for a second opinion to evaluate for symptoms that were consistent with a demyelinating process (MS). As per Dr. [NAME]’s note, it appeared that the patient had complained of some symptoms of right lower extremity weakness. She reports dragging her legs, occasionally tripping, and subsequently falling. Her exam as of that time was significant for increased reflexes in her lower extremity and subtle weakness in her right lower extremity. The working diagnosis at that time was probable multiple sclerosis, but that diagnosis was not confirmed, because we did not have the laboratory studies to confirm that diagnosis. She did have an MRI of her brain done that showed lesions that were consistent with MS, but we wanted more studies to review. The patient was subsequently lost to follow up, and as a result, we were unable to come to a diagnosis. She recently has been followed by her PCP who referred her to see a neurosurgeon given her symptoms. She underwent multiple imaging studies of her brain and total spine, and those findings were somewhat consistent with a demyelinating process, so she was referred back to our Neurology Clinic for further evaluation. Today, in clinic, she reports that she has had symptoms for over 10 years plus, and they have somewhat worsened, but not significantly. She continues to report on occasion she would drag her right foot and would sometimes trip over things. She reports difficulty with ambulation, but she seems adjusted since the symptoms. She reports chronic lower back pain that all started when she developed this lower extremity weakness, but she also reports a history of prior back trauma in a motor vehicle accident many years ago. She reports that she has some difficulty initiating ambulation after being in a seated position for an extended period of time. She reports occasional numbness in her feet (they feel heavy, and I cannot really feel them), but for most part, she has no sensory disturbances. She denies any bowel or urinary dysfunction. She denies any headaches. She denies she has ever had any visual symptoms and currently does not have any visual disturbances. She denies any shooting pain down her spine upon neck flexion. She has occasional lightheadedness when she turns her head to the left, and this immediately resolves upon repositioning. Overall, she feels like her walking is slightly worse, but not significant. She has never been on any medications that would include steroids or multiple sclerosis modulating therapies.

**IMAGING STUDY:** We were able to review the MRI of the brain and total spine that was performed in mid-2008. Her MRI showed a few tiny hyperintense lesions on the corpus callosum, a 1-cm hyperintense lesion in the left corona radiata close to left lateral ventricle, and multiple hyperintense lesions in the dorsal part of the fourth ventricle, but otherwise was normal. The MRI of the cervical spine was significant for hyperintense lesion around C3-C4 region, but otherwise no abnormality noted in the thoracic and lumbar region. None of these lesions were enhancing.

##### Gold-standard MS diagnosis status label for clinical note excerpt

An MS subspecialist independently reviewed the same patient note to determine the expected response at each decision node and the final MS diagnosis. The expected response serves as the reference standard for comparing the model outputs for this clinical note excerpt. The question at the decision node and clinical expert opinion for this clinical note excerpt are shown below.

**Diagnosis of Related Disorder Node 1: Does the patient have a diagnosis of NMO or MOGAD, or positive laboratory findings of NMO-IgG, AQP4-Ab, or MOG-Ab?** No. The patient does not have a diagnosis of NMO or MOGAD. No biomarker testing for NMO or MOGAD was performed.

**Clinical Presentation Node 1: Does the patient have at least one clinical attack?** Yes. The patient had one clinical attack in 2004. The patient reported right lower extremity weakness, dragging of legs, tripping, and falling. MRI brain is reported to have shown lesions consistent with MS. The patient continues to experience residual symptoms since this clinical attack.

**Clinical Presentation Node 2: Does the patient have MRI evidence of at least one MS lesion?** Yes. The patient has MRI evidence of at least one MS lesion. MRI brain in mid 2008 revealed one periventricular lesion and multiple infratentorial lesions. MRI spine revealed one cervical lesion.

**Dissemination in Space Node 1: Does the patient have MRI evidence of at least one MS lesion in two distinct MS typical regions (periventricular, cortical or juxtacortical, infratentorial, or spinal cord)?** Yes. The patient has MRI evidence of at least one MS lesion in three MS typical regions (periventricular, infratentorial, cervical spine). MRI brain in mid 2008 revealed one periventricular lesion and multiple infratentorial lesions. MRI spine revealed one cervical lesion.

**Dissemination in Space Node 2: Does this patient have an additional clinical attack involving a different central nervous system location than the first clinical attack?** Not applicable. Did not reach the decision node.

**Dissemination in Time Node 1: Does the patient have two confirmed clinical attacks occurring at least 30 days apart?** No. The patient note does not suggest that two clinical attacks occurred at least 30 days apart. There is evidence of one clinical attack characterized by right lower extremity weakness and difficulty with ambulation. Although the patient continues to experience residual symptoms over 10 years, there is no documentation of any additional clinical attacks.

**Dissemination in Time Node 2: Are enhancing lesions and non-enhancing lesions present together on MRI at any time?** No. The patient's MRI revealed several T2 hyperintense (non-enhancing) lesions, no enhancing lesions were present.

**Dissemination in Time Node 3: Is there evidence of a new hyperintense T2 lesion or new enhancing lesion on follow-up or repeat MRI?** No. The patient note only contains MRI results from mid 2008, no additional MRI studies were documented.

**Dissemination in Time Substitute Node 1: Is there evidence of cerebrospinal fluid (CSF)-specific oligoclonal bands?** No. CSF biomarker testing was not performed.

**MS Diagnosis Terminal Node: What is the diagnosis?** MS not confirmed (need evidence of DIT)

##### Model Performance

All models were prompted to assign a diagnosis using predefined categories. Models 1–2 generated narrative reasoning, whereas Models 3–5 incorporated the 2017 McDonald criteria decision tree and produced structured, node-based reasoning.

###### Model 1: Clinical note alone

**Predicted diagnosis:** The patient's diagnosis is Multiple Sclerosis (**MS**) confirmed.

**Rationale:** The patient has a **history of multiple clinical attacks**, including right lower extremity weakness, difficulty initiating ambulation, and occasional numbness in her feet, that have been ongoing for over 10 years. These attacks have **occurred more than 30 days apart**. The MRI evidence shows multiple lesions in the brain (corpus callosum, left corona radiata, and dorsal part of the fourth ventricle) and cervical spine (C3-C4 region), which are consistent with MS. These **lesions are in different regions of the central nervous system, demonstrating Dissemination in Space** (**DIS**). The patient's symptoms and MRI findings meet the 2017 McDonald Criteria for a confirmed diagnosis of MS.

###### Model 2: Clinical note with expert-curated knowledge and narrative description of 2017 McDonald criteria

**Predicted diagnosis:** The patient's diagnosis is MS not confirmed (lacking evidence of DIS and DIT).
**Rationale:** The patient has had symptoms consistent with a clinical attack, including right lower extremity weakness, increased reflexes in her lower extremity, and difficulty with ambulation. These **symptoms have been ongoing for over 10 years,** suggesting they meet the criteria for a duration of at least 24 hours. However, there is **no clear evidence of a second clinical attack occurring at least 30 days apart from the first,** which is necessary for confirmation of MS according to the 2017 McDonald Criteria. Additionally, while the patient's MRI shows **multiple hyperintense lesions in various locations**, including the corpus callosum and the left corona radiata, there is **no mention of enhancing lesions,** which would be indicative of an ongoing clinical attack. Therefore, both DIS and DIT are not confirmed based on the information available in this patient note.

###### Model 3: Decision tree based on 2017 McDonald criteria

**Diagnosis of Related Disorder Node 1: Does the patient have a diagnosis of NMO or MOGAD, or positive laboratory findings of NMO-IgG, AQP4-Ab, or MOG-Ab?** No. The patient note does not provide any information on a diagnosis of neuromyelitis optica (NMO) or myelin oligodendrocyte glycoprotein antibody disease (MOGAD), nor does it mention any positive laboratory findings of NMO-IgG, AQP4-Ab, or MOG-Ab. The patient's symptoms and MRI findings are suggestive of a demyelinating process, and multiple sclerosis was considered as a probable diagnosis in the past. However, without specific laboratory results or a confirmed diagnosis, it cannot be definitively stated that the patient has NMO, MOGAD, or positive findings of NMO-IgG, AQP4-Ab, or MOG-Ab.

**Clinical Presentation Node 1: Does the patient have at least one clinical attack?** Yes, the patient has had at least one clinical attack. The patient's history of symptoms such as right lower extremity weakness, dragging her legs, occasionally tripping and falling, increased reflexes in her lower extremity, and difficulty initiating ambulation after being seated for an extended period of time, all indicate a neurological event or "attack." These symptoms have been present for over 10 years and have somewhat worsened. The MRI findings of hyperintense lesions in the brain and cervical spine are also consistent with a demyelinating process such as multiple sclerosis, which typically presents with clinical attacks.

**Clinical Presentation Node 2: Does the patient have MRI evidence of at least one MS lesion?** Yes, the patient does have MRI evidence of at least one MS lesion. The MRI of the brain showed a few tiny hyperintense lesions on the corpus callosum, a 1-cm hyperintense lesion in the left corona radiata close to left lateral ventricle, and multiple hyperintense lesions in the dorsal part of the fourth ventricle. Additionally, the MRI of the cervical spine was significant for a hyperintense lesion around the C3-C4 region. These findings are consistent with MS, although further studies would be needed for a definitive diagnosis.

**Dissemination in Space Node 1: Does the patient have MRI evidence of at least one MS lesion in two distinct MS typical regions (periventricular, cortical or juxtacortical, infratentorial, or spinal cord)?** Yes, there is MRI evidence of at least one MS lesion in two distinct MS typical regions. The patient's MRI showed a few tiny hyperintense lesions on the corpus callosum (periventricular region), a 1-cm hyperintense lesion in the left corona radiata close to left lateral ventricle (periventricular region), and multiple hyperintense lesions in the dorsal part of the fourth ventricle (infratentorial region). Additionally, the MRI of the cervical spine was significant for a hyperintense lesion around the C3-C4 region (spinal cord region). These findings are consistent with the McDonald criteria for diagnosing MS, which includes dissemination of lesions in space (at least one lesion in at least two out of four MS-typical regions of the CNS: periventricular, cortical or juxtacortical, infratentorial, and spinal cord).

**Dissemination in Space Node 2: Does this patient have an additional clinical attack involving a different central nervous system location than the first clinical attack?** Not applicable. Did not reach the decision node.

**Dissemination in Time Node 1: Does the patient have two confirmed clinical attacks occurring at least 30 days apart?** No. The patient note does not provide clear evidence of at least two confirmed clinical attacks occurring at least 30 days apart. The note mentions the patient has had symptoms for over 10 years and they have somewhat worsened, but not significantly. However, it does not specify distinct episodes or attacks that could be clearly separated by a period of 30 days or more. The symptoms seem to be more of a chronic, ongoing nature rather than distinct attacks.

**Dissemination in Time Node 2: Are enhancing lesions and non-enhancing lesions present together on MRI at any time?** No. The patient note does not provide evidence of both enhancing and non-enhancing lesions present together on the MRI at any time. The note mentions multiple hyperintense lesions in various parts of the brain and spine, but it specifically states that "none of these lesions were enhancing." Therefore, based on the information available in this patient note, only non-enhancing lesions are present.

**Dissemination in Time Node 3: Is there evidence of a new hyperintense T2 lesion or new enhancing lesion on follow-up or repeat MRI?** No. The patient note does not provide evidence of a new hyperintense T2 lesion or new enhancing lesion on follow-up or repeat MRI. The note mentions the MRI results from mid 2008, which showed a few tiny hyperintense lesions on the corpus callosum, a 1-cm hyperintense lesion in the left corona radiata close to the left lateral ventricle, and multiple hyperintense lesions in the dorsal part of the fourth ventricle. However, it does not provide information on any new lesions or enhancements compared to previous MRI results.

**Dissemination in Time Substitute Node 1: Is there evidence of cerebrospinal fluid (CSF)-specific oligoclonal bands?** No. The patient note does not provide any information on the presence or absence of cerebrospinal fluid (CSF)-specific oligoclonal bands. These bands are often tested for in the diagnostic process for multiple sclerosis, as they are found in the CSF of about 85-95% of people with MS. However, the note does not mention any lumbar puncture or CSF analysis being performed, so we cannot determine whether or not this patient has CSF-specific oligoclonal bands.

**MS Diagnosis Terminal Node: What is the diagnosis?** MS not confirmed (need evidence of DIT)

###### Model 4: Decision tree with expert-curated clinical knowledge

**Diagnosis of Related Disorder Node 1: Does the patient have a diagnosis of NMO or MOGAD, or positive laboratory findings of NMO-IgG, AQP4-Ab, or MOG-Ab?** No. The patient note does not provide information on whether the patient has a diagnosis of neuromyelitis optica (NMO) or myelin oligodendrocyte glycoprotein antibody disease (MOGAD), or a positive laboratory findings of NMO-IgG, AQP4-Ab, or MOG-Ab.

The note mentions that the patient was suspected to have a demyelinating process such as MS, but no definitive diagnosis was made due to lack of laboratory studies. The note does not mention any serological testing for NMO or MOGAD specific biomarkers. Therefore, based on the information available in this patient note, it cannot be determined if the patient has NMO, MOGAD, or positive findings of NMO-IgG, AQP4-Ab, or MOG-Ab.

**Clinical Presentation Node 1: Does the patient have at least one clinical attack?** Yes, the patient has had at least one clinical attack. The patient's history of present illness indicates that she has had symptoms for over 10 years, which have somewhat worsened. She reports dragging her right foot, tripping over things, and having difficulty with ambulation. These symptoms are consistent with a clinical attack involving the cerebrum and spinal cord, as they include limb weakness and gait disturbance. The patient's symptoms are persistent and have been present for an extended period of time, meeting the criteria for a clinical attack (symptoms lasting for at least 24 hours). The patient also denies any fever, infection, stress, or heat exposure, which further supports the occurrence of a clinical attack. Furthermore, the patient's MRI findings of hyperintense lesions in the brain and cervical spine are consistent with a demyelinating process, which supports the clinical history of a neurological attack.

**Clinical Presentation Node 2: Does the patient have MRI evidence of at least one MS lesion?** Yes, the patient has MRI evidence of at least one MS lesion. The MRI of the brain showed a few tiny hyperintense lesions on the corpus callosum, a 1-cm hyperintense lesion in the left corona radiata close to left lateral ventricle, and multiple hyperintense lesions in the dorsal part of the fourth ventricle. The MRI of the cervical spine was significant for a hyperintense lesion around the C3-C4 region. These findings are consistent with the appearance of MS lesions on MRI, which are typically hyperintense on T2-weighted imaging and FLAIR sequences.

**Dissemination in Space Node 1: Does the patient have MRI evidence of at least one MS lesion in two distinct MS typical regions (periventricular, cortical or juxtacortical, infratentorial, or spinal cord)?** Yes, there is MRI evidence of at least one MS lesion in two distinct MS typical regions. The patient note mentions the presence of a few tiny hyperintense lesions on the corpus callosum, which is considered a periventricular region. Additionally, there are multiple hyperintense lesions in the dorsal part of the fourth ventricle, which is an infratentorial region. Furthermore, there is a hyperintense lesion around the C3-C4 region of the cervical spine, which is considered part of the spinal cord region. Therefore, the MRI findings indicate the presence of MS lesions in the periventricular, infratentorial, and spinal cord regions.

**Dissemination in Space Node 2: Does this patient have an additional clinical attack involving a different central nervous system location than the first clinical attack?** Not applicable. Did not reach the decision node.

**Dissemination in Time Node 1: Does the patient have two confirmed clinical attacks occurring at least 30 days apart?** No, the patient does not have at least two confirmed clinical attacks occurring at least 30 days apart. The patient has had symptoms for over 10 years, but it is not clear whether these symptoms have been continuous or whether they have occurred in distinct episodes. The patient's symptoms include right lower extremity weakness, difficulty with ambulation, and occasional numbness in her feet, but these symptoms have not been explicitly described as occurring in distinct episodes that could be classified as clinical attacks. Furthermore, the patient has not reported any symptoms that are typical of clinical attacks in multiple sclerosis, such as optic neuritis, visual loss, or acute transverse myelopathy. Therefore, based on the information provided in the patient note, it cannot be confirmed that the patient has had at least two clinical attacks occurring at least 30 days apart.

**Dissemination in Time Node 2: Are enhancing lesions and non-enhancing lesions present together on MRI at any time?** No, the patient's MRI does not show the presence of both enhancing and non-enhancing lesions together at any time. The note specifically states that "None of these lesions were enhancing." The lesions described in the MRI results, such as the few tiny hyperintense lesions on the corpus callosum, a 1-cm hyperintense lesion in the left corona radiata close to left lateral ventricle, and multiple hyperintense lesions in the dorsal part of the fourth ventricle, as well as the hyperintense lesion around C3-C4 region in the cervical spine, are all non-enhancing lesions. The absence of any mention of gadolinium-enhancing lesions or new or enlarging active lesions indicates that there were no enhancing lesions present.

**Dissemination in Time Node 3: Is there evidence of a new hyperintense T2 lesion or new enhancing lesion on follow-up or repeat MRI?** No, there is no evidence of a new hyperintense T2 lesion or new enhancing lesion on follow-up or repeat MRI. The patient note mentions an MRI that was performed in mid 2008, but it does not provide information about a follow-up or repeat MRI for comparison. The MRI findings mentioned in the note include a few tiny hyperintense lesions on the corpus callosum, a 1-cm hyperintense lesion in the left corona radiata close to left lateral ventricle, and multiple hyperintense lesions in the dorsal part of the fourth ventricle. However, without a previous MRI for comparison, it is not possible to determine if these are new lesions. Furthermore, none of these lesions were enhancing, which would indicate acute inflammation or an ongoing clinical attack.

**Dissemination in Time Substitute Node 1: Is there evidence of cerebrospinal fluid (CSF)-specific oligoclonal bands?** No. The patient note does not provide information on whether there is evidence of cerebrospinal fluid (CSF)-specific oligoclonal bands. CSF-specific oligoclonal bands are often tested for in the diagnostic process of multiple sclerosis, but this note does not mention any lumbar puncture or CSF analysis being performed.

**MS Diagnosis Terminal Node: What is the diagnosis?** MS not confirmed (need evidence of DIT)

###### Model 5: Decision tree, expert-curated clinical knowledge, and examples for few-shot learning

**Diagnosis of Related Disorder Node 1: Does the patient have a diagnosis of NMO or MOGAD, or positive laboratory findings of NMO-IgG, AQP4-Ab, or MOG-Ab?** No. There is insufficient evidence in the clinical note to address this question.

**Clinical Presentation Node 1: Does the patient have at least one clinical attack?** Yes, the patient has had at least one clinical attack. The patient reported right lower extremity weakness and unsteady gait that has persisted for over 10 years. These symptoms meet the criteria for a clinical attack as they involve the central nervous system, have lasted for more than 24 hours, and occurred in the absence of fever, infection, stress, or heat exposure. The patient's symptoms prompted clinical evaluation and imaging, further supporting the classification of these symptoms as a clinical attack. The MRI findings showing lesions consistent with MS further support this conclusion.

**Clinical Presentation Node 2: Does the patient have MRI evidence of at least one MS lesion?** Yes, the patient has MRI evidence of at least one MS lesion. The MRI showed a few tiny hyperintense lesions on the corpus callosum, a 1-cm hyperintense lesion in the left corona radiata close to left lateral ventricle, and multiple hyperintense lesions in the dorsal part of the fourth ventricle. Additionally, the MRI of the cervical spine was significant for a hyperintense lesion around the C3-C4 region. These findings are consistent with demyelinating lesions associated with multiple sclerosis.

**Dissemination in Space Node 1: Does the patient have MRI evidence of at least one MS lesion in two distinct MS typical regions (periventricular, cortical or juxtacortical, infratentorial, or spinal cord)?** Yes, the MRI findings in the patient note indicate the presence of MS lesions in two distinct MS typical regions. Specifically, hyperintense lesions were found in the periventricular region (corpus callosum and left corona radiata close to left lateral ventricle) and in the cervical spinal cord (C3-C4 region). These findings meet the criteria for dissemination in space according to the McDonald criteria for MS diagnosis.

**Dissemination in Space Node 2: Does this patient have an additional clinical attack involving a different central nervous system location than the first clinical attack?** Not applicable. Did not reach the decision node.

**Dissemination in Time Node 1: Does the patient have two confirmed clinical attacks occurring at least 30 days apart?** Yes. The patient has had at least two confirmed clinical attacks occurring at least 30 days apart. The first attack occurred more than 10 years ago, when the patient first noticed symptoms of right lower extremity weakness, dragging her leg, tripping, and falling. The second attack occurred in 2004, when the patient was seen in the clinic and was found to have increased reflexes in her lower extremity and subtle weakness in her right lower extremity. These attacks are separated by a period of several years, far exceeding the 30-day requirement. Both attacks involved the patient's right lower extremity, suggesting a possible focal lesion in the central nervous system. The patient's symptoms lasted for more than 24 hours and occurred in the absence of fever, infection, stress, or heat exposure, meeting the criteria for a clinical attack.

**Dissemination in Time Node 2: Are enhancing lesions and non-enhancing lesions present together on MRI at any time?** Not applicable. Did not reach the decision node.

**Dissemination in Time Node 3: Is there evidence of a new hyperintense T2 lesion or new enhancing lesion on follow-up or repeat MRI?** Not applicable. Did not reach the decision node.

**Dissemination in Time Substitute Node 1: Is there evidence of cerebrospinal fluid (CSF)-specific oligoclonal bands?** Not applicable. Did not reach the decision node.

**MS Diagnosis Terminal Node: What is the diagnosis?** MS confirmed

##### Findings of Iterative Model Development

Across the subset of three clinically complex notes, the decision tree with clinical knowledge (Model 4) showed the most consistent agreement with expert-determined reference standard responses and diagnoses, and was selected for the primary analysis.

#### SUPPLEMENTARY TABLES

#### S-Table 1. Examples of GPT-4 hallucinations during decision tree traversal.

We illustrate hallucination types using representative examples. For each, we include relevant clinical note excerpts, the decision node prompt, GPT-4’s response, and clinical expert opinion. Hallucination categories include non-factual (output contradicts facts), incoherent (output contradicts input context or dialogue is self-contradictory), irrelevant (output is unrelated to the topic), overreliant (excessive reliance on provided context without applying provided knowledge), or logical reasoning errors (error in logic or understanding). Categories were assigned by manual chart review, and responses could exhibit multiple hallucination types.

| **Type of Hallucination** | **Excerpt from Clinical Note** | **Decision Node** | **GPT-4 Response** | **Clinical Expert Opinion** |
| --- | --- | --- | --- | --- |
| Non-factual | MRI brain with and without contrast revealed an approximately 4mm focus just posterior to the left atrium and 2-3mm area of signal abnormality is present within the deep white matter at the lateral aspect of the left frontal lobe. MRI cervical with and without contrast: Area of abnormal T2 signal noted at level C6 in the cervical spinal cord. | Dissemination in Space Node 1:  Is there MRI evidence of at least one lesion in two distinct MS-specific regions (periventricular, cortical or juxtacortical, infratentorial, or spinal cord)? | No lesions in ≥2 MS-typical regions.  Brain MRI findings reveal two areas of **white matter signal abnormality: one posterior to the left atrium** and another in the deep white matter at the lateral aspect of the left frontal lobe. **These lesions are not in MS-specific regions**. | Disagree.  Lesion posterior to the left atrium should be classified as periventricular, an MS-typical region. |
| Incoherent | Cranial MRI which showed 1 pericallosal region signal anomaly and 2 larger periventricular signal anomalies. There was also an area of increased enhancement in the basal ganglia. | Dissemination in Time Node 2:  Are enhancing lesions and non-enhancing lesions present together on MRI at any time? | No enhancing + non-enhancing lesions.  The note **does not explicitly state that there are both enhancing and nonenhancing lesions** present at the same time. The note only mentions one area of enhancement (basal ganglia). | Disagree.  Pericallosal and periventricular signal anomalies should be categorized as non-enhancing lesions by default. |
| Irrelevant | Cranial MRI showed 1 pericallosal region signal anomaly and 2 larger periventricular signal anomalies. There was also an area of increased enhancement in the basal ganglia. | Clinical Presentation Node 2:  Does the patient have MRI evidence of at least one MS lesion? | Yes, ≥1 MS lesion.  The patient's MRI showed 1 pericallosal region signal anomaly as well as 2 larger periventricular signal anomalies.  **The patient's symptoms, including difficulty speaking, problems with balance, decline in handwriting skills are also consistent with MS.** | Agree, but irrelevant.  Symptom information was not pertinent to the lesion-focused question. |
| Overreliant | The examination demonstrates several scattered foci of abnormal T2 prolongation and density elevation in a periventricular distribution and in the deep white matter of the centrum semiovale. | Dissemination in Space Node 1:  Is there MRI evidence of at least one lesion in two distinct MS specific regions (periventricular, cortical or juxtacortical, infratentorial, or spinal cord)? | No lesions in ≥2 MS-typical regions.  While **periventricular** and deep white matter **changes** are mentioned, these are **not specifically identified as MS lesions.** | Disagree.  Periventricular abnormalities were not classified as MS lesions due to lack of explicit labeling. |
| Logical reasoning | [PATIENT NAME] had an episode of numbness and tingling (on the right side of face, periorbital / perioral and cheek, sparing forehead, lasting 6 hours). | Dissemination in Time Node 1:  Does the patient have at least two confirmed clinical attacks that involve different symptoms occurring at least 30 days apart? | No <2 confirmed clinical attacks.  The patient had an episode of numbness and tingling on the right side of her face that **lasted for 6 hours.** Although this episode did not last for more than 24 hours, it **prompted clinical evaluation, thus meeting the criteria for a clinical attack.** | Disagree.  Symptoms lasting 6 hours do not meet criteria for a clinical attack, regardless of clinical evaluation. |

#### SUPPLEMENTARY FIGURES

#### S-Figure 1. Example of node-level model reasoning for clinical attack determination.

We illustrate an example of node-level reasoning during decision tree traversal. The clinical note excerpt, corresponding prompt, GPT-4 response, and clinical expert interpretation are shown. Text highlighting denotes positive (green) and negative (red) clinical features informing the decision. The expert interpretation reflects the reference standard for the expected response at this node.

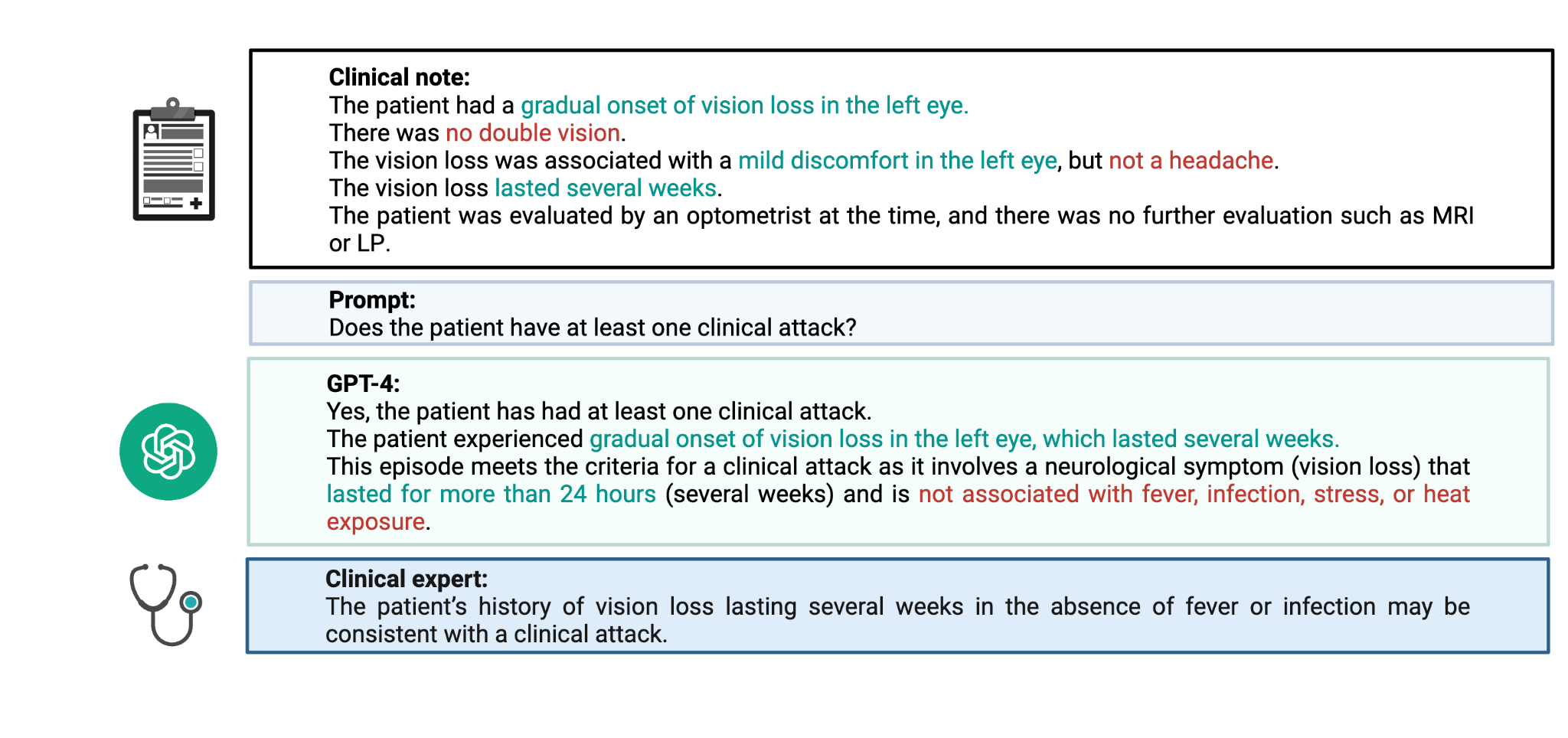

#### S-Figure 2. Illustrative example of node-level model reasoning with incoherent and irrelevant hallucinations.

The figure illustrates an example of model reasoning at a single decision node. Shown are the clinical note excerpt, node-specific prompt, GPT-4 response, and clinical expert evaluation. Text highlighting denotes relevant (green) and non-relevant (red) clinical features with respect to the prompt. The model failed to identify the enhancing basal ganglia lesion, an MS-typical site defined in the curated knowledge (incoherence hallucination), and included symptom information not pertinent to the lesion-focused query (irrelevance hallucination).

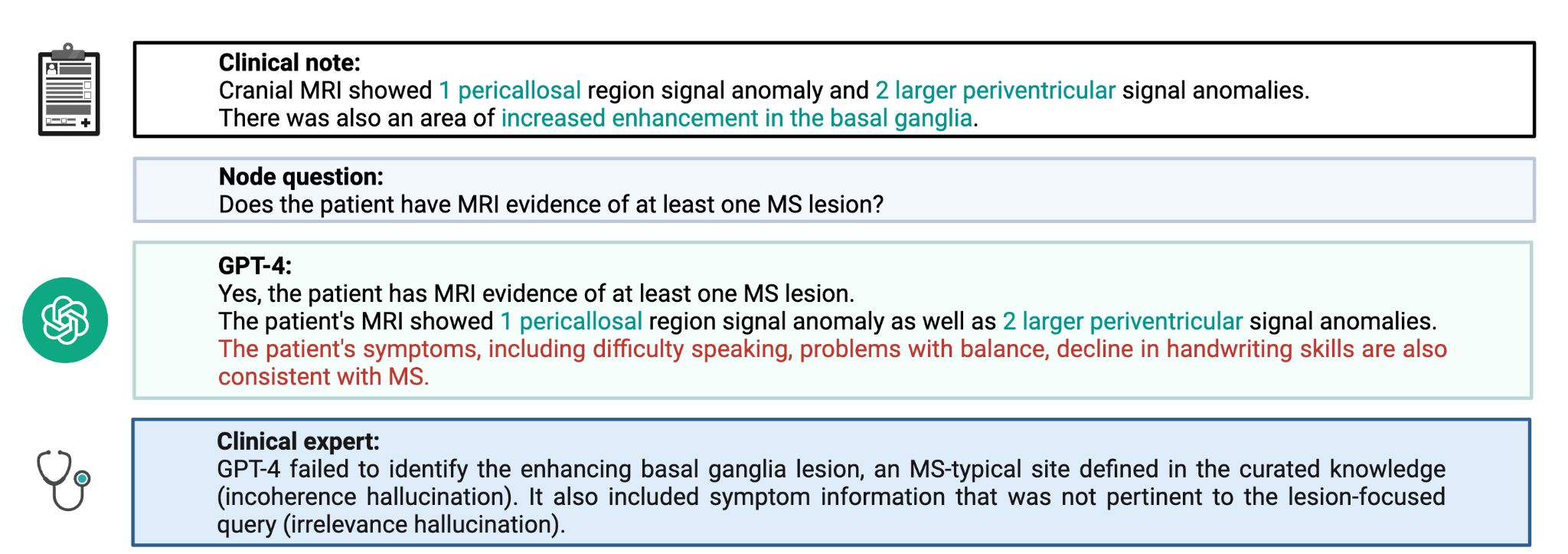

#### S-Figure 3. Comparison of Model Configurations for MS Diagnosis Derivation.

Five model configurations were used to derive MS diagnoses from clinical documentation, differing in the degree of structured criteria, clinical knowledge, and example-based guidance provided to the model.

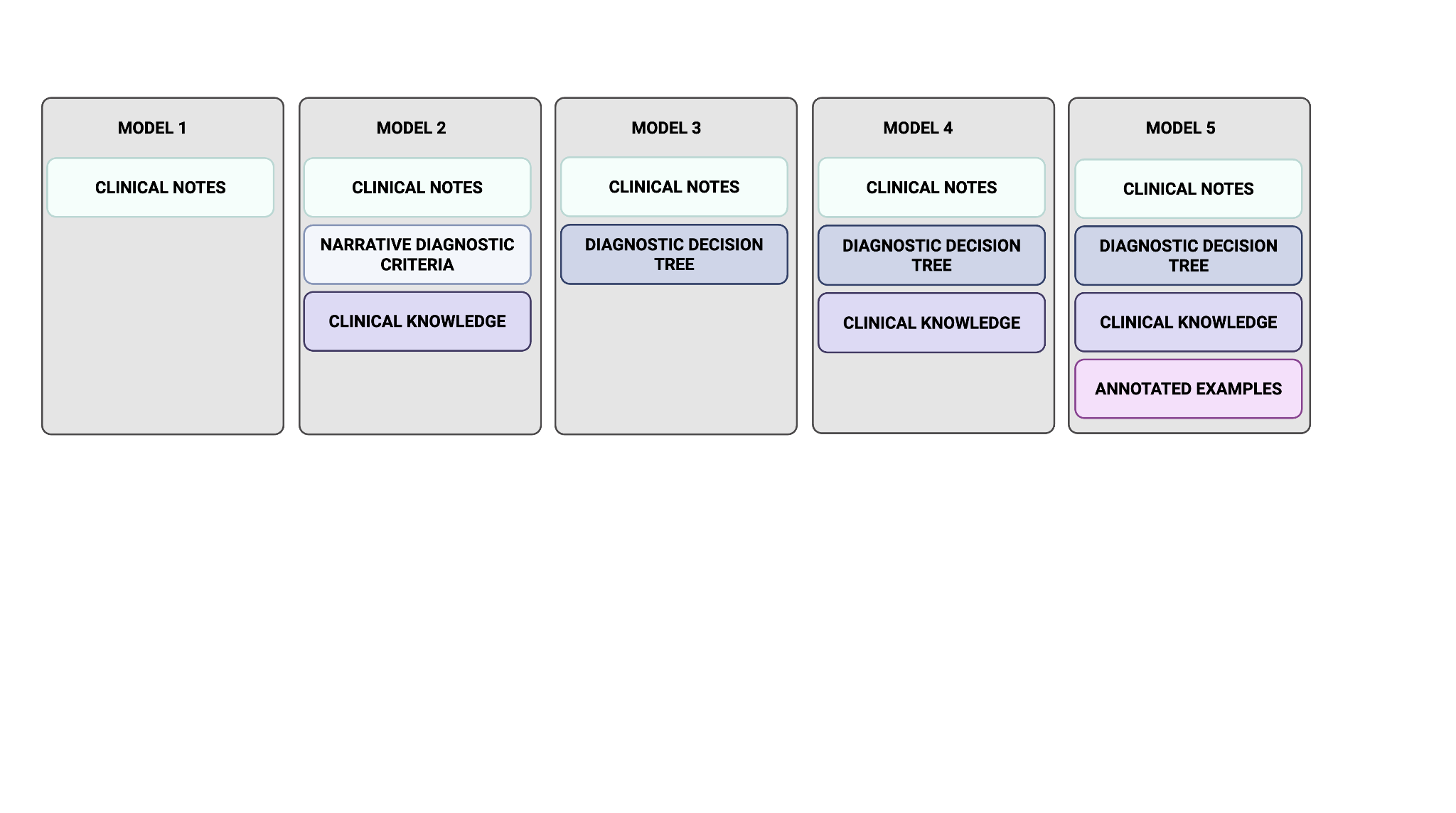
